# No evidence that retinol is protective for skin cancer

**DOI:** 10.1101/2024.08.27.24312670

**Authors:** Marloes Helder, Nirmala Pandeya, Mathias Seviiri, Catherine M. Olsen, David C. Whiteman, Matthew H. Law

## Abstract

With over 1.5 million new cases annually, skin cancers are the most commonly diagnosed group of cancers worldwide. Among these, melanoma and keratinocyte cancers (KC), comprising squamous cell carcinoma (SCC) and basal cell carcinoma (BCC), are predominant. Retinol, a vitamin A derivative, is essential in the regulation of growth and differentiation of epidermal cells. Moreover, retinol exhibits antioxidant properties, protecting the skin against ultra-violet (UV) radiation induced oxidative damage.

Existing research on the impact of retinol on melanoma, SCC and BCC development shows mixed results. Several dietary intake studies have suggested that higher retinol levels reduce skin cancer risk, however, others have failed to find this association.

We used two-sample Mendelian randomization (MR) to explore if there is a causal relationship between retinol and the risk of developing melanoma, SCC or BCC. Genetically predicted circulating retinol levels were obtained from a genome wide association study (GWAS) meta-analysis of the INTERVAL (N=11,132) and METSIM (N=6,136) cohorts. Melanoma (30,134 cases and 375,188 controls), SCC (10,557 cases and 537,850 controls) and BCC (36,479 cases and 540,185 controls) risks were derived from published GWAS meta-analyses. We conducted two MR approaches. In the first MR we used a single SNP (rs10882283) that is associated with the levels of Retinol Binding Protein 4 (RBP4) as an instrument variable (IV) for circulating retinol levels. In the second MR we used all independent genetic variants that were strongly associated (P < 5 × 10^−8^) with retinol levels as IVs. Odds ratios (OR) for skin cancer were calculated for a one standard deviation (SD) increase in genetically predicted retinol levels.

The single IV approach revealed that retinol levels were not significantly associated with risk of melanoma (OR = 1.04 [95% confidence interval 0.83, 1.31], P = 0.72), SCC (OR = 1.15[0.87, 1.51], P = 0.32) or BCC (OR = 1.06 [0.90, 1.23], P = 0.50). Similar null results were observed with the multiple IV approach for melanoma (OR = 1.03 [0.95, 1.11], P = 0.54), SCC (OR = 1.01 [0.91, 1.13], P = 0.83), and BCC (OR = 1.04 [0.96, 1.12], P = 0.38).

In conclusion, we found no evidence that circulating retinol levels were causally associated with the development of melanoma, SCC and BCC.

## INTRODUCTION

Skin cancer is the most commonly diagnosed group of cancers worldwide, with Australia leading with the highest rates overall^1^. Particularly within fair-skinned populations of European ancestry, incidence rates have increased over the last 50 years, with projections indicating a further increase by more than 50% by 2040^2,3^. A major challenge for KC research is that most cancer registries do not capture diagnosis of KC due to their very high incidence and low mortality^2–4^. Nonetheless, KC imposes a substantial burden on patients due to their direct morbidity and the impact of treatment, such as surgical excision or radiotherapy^5,6^. Moreover, skin cancer also exacts a heavy financial toll, with the Australian health system expending over 1.5 billion AUD per year, which is expected to increase in the coming years^6^.

While ultra-violet (UV) radiation exposure and fair-skin are well-established risks, numerous other factors have been associated with skin cancer. There has been a considerable interest in the concept of photoprotection through dietary antioxidants. Several micronutrients have the potential to reduce the genotoxic properties of reactive oxygen species^7,8^. However, observational and dietary recall studies can be impacted by a range of biases, making it difficult to determine if any observed associations are causal. It is possible however to use Mendelian randomisation to explore if observed associated are consistent with a causal association. MR uses genetic variation to explore causal relationships between an exposure (in this case dietary oxidants) and outcome (here skin cancer). Causality can be inferred as genetic variants associated with exposure are inherited at conception and are independent of other potential confounders. For instance, serum 25-hydroxyvitamin D has been linked in observational studies to an increased risk for melanoma and KC, yet recent MR studies have found that lower 25-hydroxyvitamin D is unlikely to be a causal risk factor for skin cancer^9,10^.

Another skin-related candidate compound is retinol, an active form of vitamin A present in animal-derived foods. Retinol is essential in the regulation of the growth and differentiation of epidermal cells and the stimulation of collagen synthesis^11^. Furthermore, retinol has antioxidant properties, offering protection against UV-radiation induced genotoxicity^11^. These mechanisms suggest a potential protective role of retinol in skin cancer development. Previous studies that used dietary intake as a measure for retinol exposure yielded mixed results for skin cancer. For instance, one study found a small but significant protective association between higher dietary retinol intake and SCC risk^12^. However, other studies failed to find this association^13^. It is important to note that there are different approaches to infer possible associations between retinol and skin cancer, from epidemiological studies that measure intake based on dietary recall, through to experimental studies that measure serum retinol levels directly, and clinical trials that assign individuals to receive retinol. These different study designs must be considered when comparing results, as each approach can have different confounders and biases.

A recent large genome-wide association study (GWAS) of circulating retinol levels investigated retinol’s association with melanoma and KC using MR^14^. While no significant results were observed for melanoma (P = 0.76), or SCC and BCC together (P = 0.24), these results were underpowered due to the modest number of skin cancer cases, particularly for melanoma and SCC. Moreover, the GWAS used for the outcome did not separate BCC and SCC, precluding separate analyses for these cancers. We sought to explore the relationship between circulating retinol levels and the different types of skin cancer (melanoma, SCC and BCC) in more detail using well-powered disease-specific GWAS.

## METHODS

### Study cohort

Summary statistics for genetic variants associated with circulating retinol were drawn from the inverse-variance weighted (IVW) GWAS meta-analysis conducted by Reay et al^14^. The details of this GWAS are outlined in the Reay et al. paper. Briefly, the INTERVAL and METSIM cohorts were included, with 11,132 and 6,136 participants of European ancestry respectively. The INTERVAL study, with recruitment from 2012 to 2014, consists of adult blood donors from the United Kingdom^15^. The METSIM cohort, with baseline visits from 2005 and 2010, consists of middle-aged men from Northern Finland^16^. Summary statistics for genetic variants associated with melanoma were from the confirmed-cases only GWAS meta-analysis of Landi et al., consisting of 30,134 cases and 375,188 controls of European ancestry^17^. Further information regarding the studies included and their recruitment process is available in the paper^17^. We drew summary statistics for genetic variants associated with BCC and SCC from the GWAS meta-analysis reported in Keatley et al^18^. This meta-analysis included data from QSKIN^19^, FinGen^16^ and the UK Biobank^20^, with in total 36,479 cases and 540,185 controls for BCC and 10,557 cases and 537,850 controls for SCC, all of European ancestry^18^. Ethical approval and oversight was provided by the QIMR Berghofer Human Research Ethics Committee. Each GWAS was restricted to European ancestry, based on genetic principal component analysis (PCA)^14,17,18^. Residual ancestry stratification in this study was accounted for by fitting the first 10 principal component scores.

### Selection of instrumental variables

We converted summary statistics from the GWAS data of retinol from GRCh38 to GRCh37 and harmonized the four datasets to a common set of SNPs to make sure that all SNPs of retinol were present for the three different outcomes (melanoma, SCC and BCC). We used FUMA v1.4.1 (Functional Mapping and Annotation of Genome-Wide Association Studies) platform to identify independent (linkage disequilibrium r^2^ < 0.05) SNPs robustly (P < 5 × 10^−8^) associated with circulating retinol levels^21^. Lead SNPs identified by FUMA were compared with the findings of Reay et al to ensure consistency of our analyses^14^. Because allele frequencies were not available, we replaced ambiguous (A/T or C/G) SNPs with alternative non-ambiguous SNPs exhibiting high LD.

### Statistical analyses

We conducted a series of two-sample MR analyses to assess the relationship between retinol with melanoma, SCC and BCC using the IVW method. For genetically predicted retinol levels, we used the fixed-effects meta-analysis results, with effect size units for a one standard-deviation change in retinol levels^14^. Skin cancer odds ratios (ORs) were log transformed prior to analysis. For each skin cancer type, two different instruments were used. In the first approach we used only a single SNP with a clear functional relationship with retinol levels as IV (with a fixed-effect model) to proxy circulating retinol. This approach reduces the risk of confounding or horizontal pleiotropy. In the second approach we used all genome-wide significant SNPs (with a random-effects model) as IVs to proxy retinol. This approach enhances statistical power by combining the effects of multiple SNPs, at the risk of introducing pleiotropic variants^14^. After the MR analyses, the log(OR) outcomes were converted back to OR. The *MendelianRandomization* (v0.9.0) package in Rstudio (v4.3.1) was used to perform the two-sample MR analyses^22^.

## RESULTS

After filtering, we identified nine independent lead SNPs as associated with circulating serum retinol at the genome-wide significant level of P < 5 × 10^−8^. Compared to the lead SNPs reported by Reay et al^14^, an additional SNP (rs2207132) was included. Despite rs2207132’s proximity to rs6029188, these SNPs are not in LD (r^2^ < 0.02) and both were retained. As previously reported, the strongest SNP rs10882283 is in the 5’ untranslated region of the *retinol binding protein 4* (*RBP4*) and is associated with RBP4 protein levels^23,24^. RBP4 is responsible for transporting retinol in the blood. The second strongest SNP rs1667226 is in an intron of the *transthyretin* (*TTR*) gene which encodes a carrier protein that binds to a range of transport proteins including RBP4. The first approach, the *RBP4* SNP was used as a single IV with a direct functional relationship with retinol levels (Figure 1). The single IV approach yielded an OR of 1.04 per standard deviation (SD) in genetically predicted retinol levels [95% confidence interval = 0.83, 1.31] P = 0.72 for melanoma, 1.15 [0.87, 1.51] P = 0.32 for SCC and 1.06 [0.90, 1.23] P = 0.50 for BCC. The multiple IV random-effects approach (Figure 2) revealed an OR of 1.03 [0.95, 1.11] P = 0.54 for melanoma, 1.01 [0.91, 1.13] P =0.83 for SCC and 1.04 [0.96, 1.12] P = 0.38 for BCC. All F-statistics for the individual instrumental variables were well above the threshold of 10, ruling out weak instrument bias.

**Figure 1.**
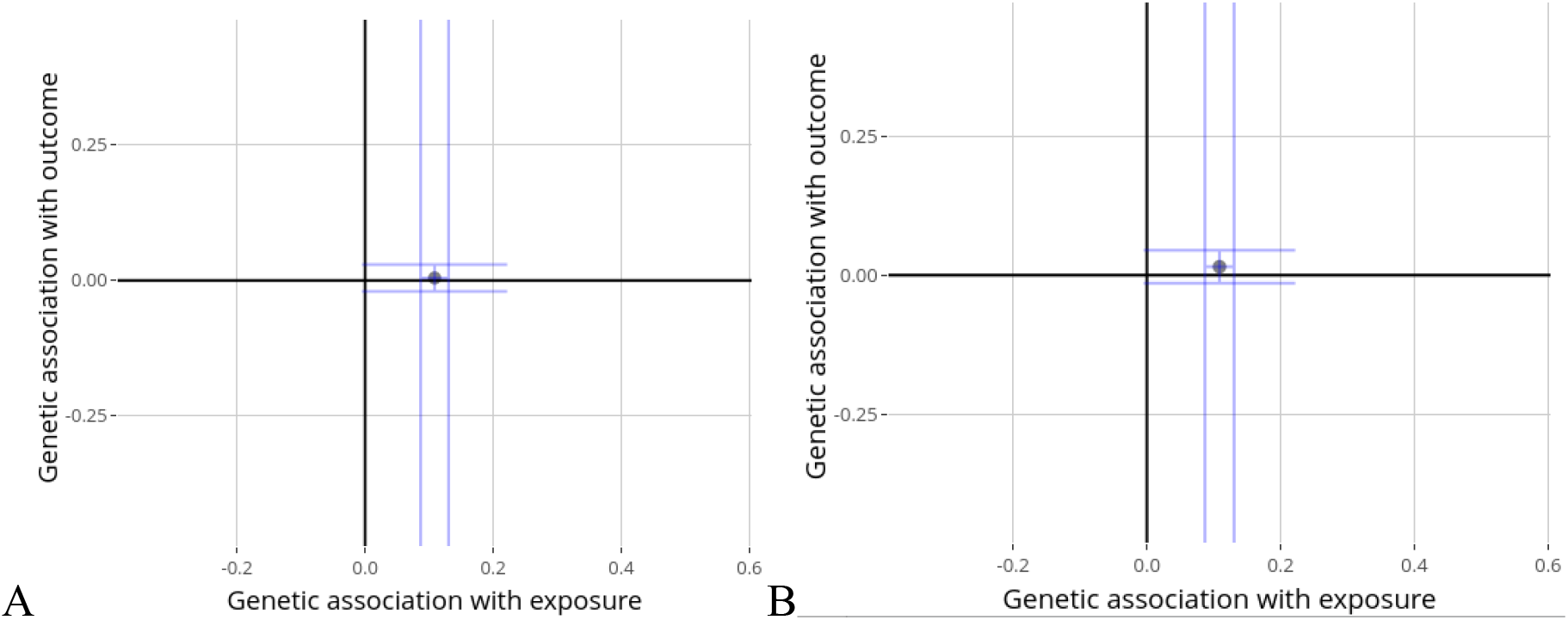
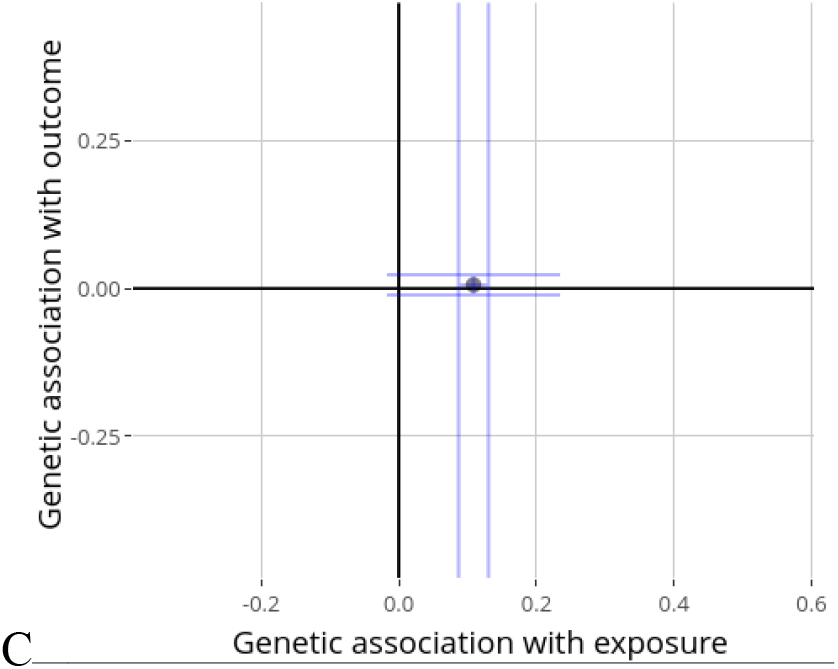
Mendelian Randomization single IV approach with rs10882283, fixed-effect model. Purple dot displays effect size estimate, blue lines the confidence interval. (A) The melanoma: log(OR) effect size estimate for a one SD change in retinol levels was 0.041 [95% confidence interval -0.19, 0.27] (P = 0.72). (B) SCC: effect size estimate of 0.14 [-0.14, 0.41](P = 0.32). (C) BCC: effect size estimate of 0.054 [-0.10, 0.21] (P = 0.50).

**Figure 2.**
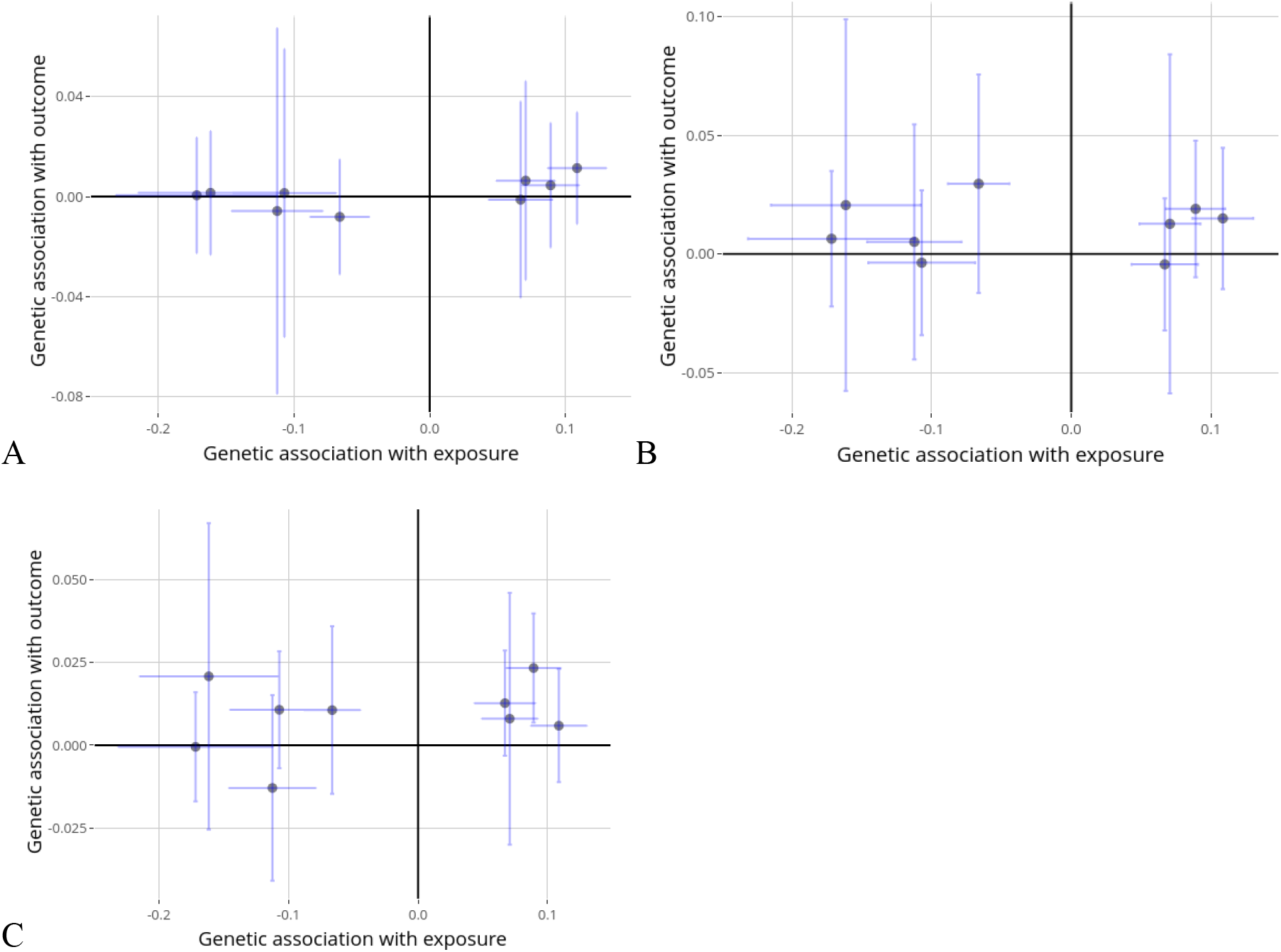
Mendelian Randomization multiple IV approach with rs11762406, rs11865979, rs1260326, rs1791209, rs2207132, rs34898035, rs4841132, rs6029188, and rs10882283 random-effect model. Purple dot displays effect size estimate, blue lines the confidence interval. (A) The melanoma: log(OR) effect size estimate for a one SD change in retinol levels was 0.025 [95% confidence interval -0.055, 0.11] (P = 0.54). (B) SCC: effect size estimate of 0.012 [-0.094, 0.12] (P = 0.83). (C) BCC: effect size estimate of 0.035 [-0.043,0.11] (P = 0.38).

**Table 1.** Summary of included SNPs in the two-sample Mendelian randomization. Effect size is a beta for retinol, and odds ratio for melanoma, SCC and BCC.

| SNP | CHR | BP | EA | NEA | Effect size |  |  |  |
| --- | --- | --- | --- | --- | --- | --- | --- | --- |
|  |  |  |  |  | Retinol | Melanoma | SCC | BCC |
| rs11762406 | 7 | 114180292 | A | C | -0.11 | 1.00 | 1.01 | 0.99 |
| rs11865979 | 16 | 79722913 | T | C | 0.067 | 1.00 | 1.00 | 1.01 |
| rs1260326 | 2 | 27730940 | T | C | 0.071 | 1.03 | 1.01 | 1.00 |
| rs1791209 | 18 | 29142754 | T | G | 0.089 | 1.00 | 1.00 | 1.01 |
| rs2207132 | 20 | 39142516 | A | G | -0.16 | 0.99 | 1.01 | 1.01 |
| rs34898035 | 2 | 122084285 | A | G | -0.17 | 1.00 | 1.02 | 1.02 |
| rs4841132 | 8 | 9183596 | A | G | -0.11 | 1.01 | 1.03 | 1.01 |
| rs6029188 | 20 | 39200914 | A | G | -0.07 | 0.98 | 1.02 | 1.02 |
| rs10882283* | 10 | 95360964 | A | C | 0.11 | 1.01 | 1.02 | 1.01 |

\*Also used in the single-instrument two-sample MR.

## DISCUSSION

Prior research has suggested that retinol may protect against certain types of skin cancer^12,25^. A recent study identified genetic variants associated with plasma retinol levels, and tested if they were causally related to a range of traits^14^. While this included GWAS for melanoma, BCC, and SCC, the individual datasets from UK Biobank and FinnGen were either underpowered, did not separate BCC and SCC, or both. We sought to more fully explore this using access to large GWAS datasets specific to melanoma, SCC and BCC. Given the robust statistical power and control of potential sources of biases (e.g. population stratification), these results suggest that it is unlikely that circulating retinol influences the risk of developing melanoma, SCC or BCC (P > 0.3). This study has sufficient power to detect small differences, with our sample size allowing us to detect an OR of 1.116 for a 1 SD change in retinol levels for melanoma, 1.105 for BCC, and 1.190 for SCC. Although we cannot rule out smaller effects, it is highly unlikely that they would be clinically significant.

The complexity of the retinol homoeostasis could be an explanation for this result. For instance, the IV used in this study correlates with circulating retinol through RBP4, which is not just specific to the skin but transports retinol to a variety of tissues via the bloodstream^26^. Another example is the distinct functions of other vitamin A derivatives besides retinol^11,27^. Once absorbed by skin cells, retinol is converted to retinoic acid (RA); excess retinoids are stored as retinyl esters to prevent RA toxicity^28^. RA subsequently activates downstream pathways regulating gene expression and various cellular processes in the skin, such as inhibiting cell growth by activating retinoic acid receptors^11^. However, RA is also mediated by alternative pathways, involving the promotion of cell survival and hyperplasia^29^. The exact cellular and molecular mechanisms by which these derivatives operate are not yet fully understood^11^.

A limitation of this study is the potential for horizontal pleiotropy when using Mendelian Randomization. We checked each SNP for associated traits that could influence both retinol and skin cancer. For SNP rs1260326 cholesterol levels appeared to be associated with both exposure and outcome, but it did not show deviating outcomes in the multiple IV approach (Figure 2). Nonetheless, by the consistency of the outcomes from the single IV with the multiple IV approach, it can be concluded that horizontal pleiotropy is probably not an issue.

Synthetically created retinoic acids demonstrate greater efficacy in inhibiting the proliferation of skin cancer cells compared to natural vitamin A derivatives^5,25,30^. Understanding the distinct mechanisms by which these synthetic variants target tumour cells may enhance our understanding of the different pathways through which retinol influences the skin. We conclude, on the basis of the best available evidence, that naturally occurring retinol shows no effect on the development of skin cancer.

## Data Availability

All data produced in the present work are contained in the manuscript.

https://zenodo.org/records/7905523

https://www.ncbi.nlm.nih.gov/projects/gap/cgi-bin/study.cgi?study_id=phs001868.v1.p1

## Acknowledgements

We acknowledge contribution of data from the retinol meta-analysis GWAS from Raey et al^14^, data from the melanoma meta-analysis consortium from Landi et al ^17^, and data from the melanoma meta-analysis GWAS from Keatley et al^18^. Please see the supplemental for full acknowledgments from contributing studies.

## Data availability

All data used in this study is available from the respective cited publications.

